# Neuropathologic and proteomic analysis of human hippocampal CA1 pyramidal neuron sublayers in human Alzheimer’s disease

**DOI:** 10.1101/2024.08.27.24312663

**Authors:** Isabel Reyes, Evgeny Kanshin, Chengju Tian, Arline Faustin, Beatrix M. Ueberheide, Arjun V. Masurkar

**Affiliations:** NYU Alzheimer’s Disease Research Center, NYU Grossman School of Medicine, New York, NY, 10016, USA; Department of Neurology, NYU Grossman School of Medicine, New York, NY, 10016, USA; Department of Biochemistry and Molecular Pharmacology, NYU Grossman School of Medicine, New York, NY, 10016, USA; Department of Pathology, NYU Grossman School of Medicine, New York, NY, 10016, USA; Neuroscience Institute, NYU Grossman School of Medicine, New York, NY, 10016, USA

**Author notes:** **Corresponding Author:** Arjun V. Masurkar, MD, PhD, NYU Grossman School of Medicine, 435 East 30th Street, 10th Floor, New York, NY 10016.

## Abstract

Hippocampal area CA1 is vulnerable to Alzheimer’s disease (AD) pathology, but if and how CA1 pyramidal neuron (PN) subpopulations are impacted is unclear. Specifically, CA1 PNs can be divided into a superficial (sPN) and deep (dPN) layer that have distinct molecular and functional properties. Understanding AD pathophysiology at the level of these CA1 PN subpopulations may inform on mechanisms of vulnerability and cognitive dysfunction. We first quantified amyloid plaque and tau pathology in Braak stage V-VI human AD cases, revealing that the sPN layer shows much higher levels of AD pathology compared to the dPN layer. We then performed quantitative localized proteomics of sPN and dPN layers in Braak stage VI human AD cases, which revealed that the two layers show disease- and function-related molecular differences that support relative hyperexcitability of sPNs and markers of delayed neurodegeneration in dPNs. In sum, these findings support that sPNs are differentially vulnerable in human AD.

## Introduction

The hippocampus is highly vulnerable to Alzheimer’s disease (AD), with its area CA1 the most affected sector ^1,2^. Even within CA1, it has been noted that amyloid plaque deposition and tau-based neurofibrillary tangle (NFT) development in pyramidal neurons (PNs) is more extensive towards subiculum (distal CA1) as opposed to near CA2 (proximal CA1) ^2^. While the mechanism of this proximo-distal gradient is unclear, CA1 PNs are known to be molecularly and functionally distinct across this same axis ^3^, which may play a role. In particular, distal and proximal CA1 are differentially driven by cortical input, with distal CA1 more strongly targeted by lateral entorhinal cortex (LEC) and proximal CA1 by medial entorhinal cortex (MEC) ^4^. As such, distal CA1 may be more susceptible to propagation of AD pathology given the early vulnerability of LEC ^2,5^.

CA1 PNs are also heterogeneous across the radial axis and can be divided into superficial and deep layers (sPNs, dPNs) that also show unique molecular, circuit, and memory-related functional properties ^3^. Yet it is unclear if these differences associate with a differential expression and impact of AD pathology on these two layers. Of note, within distal CA1, LEC preferentially excites sPNs over dPNs ^4^. In contrast, dPNs show higher intrinsic excitability ^6^. Whether these or other features associate with a differential burden of amyloid plaque and NFTs, or other molecular signatures of compromise, is unknown. Clarifying this would help improve our mechanistic understanding CA1 PN dysfunction in AD as well as mechanisms of susceptibility to AD pathology.

As such, in this study, we examined the impact of AD on CA1 sPNs and dPNs in post-mortem tissue of AD cases via two levels of analysis. First we quantified amyloid plaque and NFT burden across the two sublayers, as well as across the proximo-distal axis, to determine if AD pathology was differentially present based on somatic depth. Second, we compared the proteomic profiles of CA1 sPNs and dPNs in AD vases. While there have been prior studies examining proteomic changes in CA1 ^7-9^, these studies have not focused on differences between PN layers that may indicate differential function or differential vulnerability to AD. To accomplish this, we applied a method that combines laser capture microdissection (LCM) and quantitative liquid chromatography-mass spectrometry (LC-MS) to enable localized quantitative proteomics of these neuronal subpopulations in fixed tissue ^10-13^. As it had not been applied to CA1 PNs previously, we evaluated whether this method could robustly identify function- and disease-related peptides in human CA1 PNs and if it identified functional and disease-related differences between these layers.

## Methods

This study utilized deidentified data and samples from the NYU Alzheimer’s Disease Research Center (ADRC). The NYU ADRC operates under a protocol approved by the Institutional Review Board of the NYU Grossman School of Medicine. The NYU ADRC is a longitudinal study of aging that recruits community-dwelling older adults for annual evaluation. Inclusion criteria included age > 40, informant, fluency of participant and informant in English or Spanish, and willingness to have neuroimaging and phlebotomy performed. Exclusion criteria at entry included major neurological conditions other than AD or related dementias, and other major medical and psychiatric conditions that could otherwise impact cognition. Annual evaluations were performed according to the National Alzheimer’s Coordinating Center’s Uniform Data Set (NACC UDS) that included medical history, physical exam, surveys of cognitive and psychiatric symptoms, psychometric testing, clinical labs, and neuroimaging that informed a consensus diagnosis. A subset of participants consented to brain donation, which were processed and diagnosed per NACC UDS criteria by the NYU Neuropathology Core and stored in the NYU ADRC Brain Bank.

### Quantification of amyloid plaques and NFTs in human CA1 sPN and dPN layers

A series of 10 cases from the NYU ADRC Brain Bank were selected for analyses. In these cases, pre-prepared slides of hippocampal sections were available that had been stained with Bielschowsky stain to visualize amyloid plaques and NFTs. Slides were digitized using a conventional light microscope coupled to a digital camera, acquiring and tiling images at 60X. For analysis, 0.25mm^2^ rectangular regions of interest (ROIs) were placed around sPN and dPN layers of distal CA1. The number of amyloid plaques and NFTs were counted, as well as total cell counts in each ROI. Statistical comparisons were made using Students T-test.

### Neuronal extraction for proteomics

Human FFPE brain blocks were obtained from the NYU ADRC Brain Bank and underwent the same processes as previously described ^10^. Briefly, the brain sections of interest underwent molecular sectioning to reduce cross-contamination between samples and potential exposure to exogenous nucleases and proteases. Tissues were sectioned at 8 μm and mounted on LMD PET Steel FrameSlides (Leica Microsystems, #11505151). Twenty-four hours post-sectioning, tissues were deparaffinized and stained with cresyl violet to visualize neurons in CA1. Sections were allowed to dry for 24h prior to microdissection. Low-retention 0.5 ml microcentrifuge tubes (Axygen, #14-222-298) were loaded onto the LCM microscope (Leica, LMD6500) with 70 ul of LC/MS grade water (Fisher Scientific, #W6-4) on the cap for tissue collection. Rectangular ROIs were microdissected along the CA1 pyramidal layer at 20x. A total of 10 mm2 of tissue per sample were collected. Once the laser capture was complete, collection tubes were centrifuged at 14,000×g for 2 minutes and stored at -80 C until all samples were ready for analysis.

### Proteomics sample preparation

Samples were solubilized in the lysis buffer containing 5% (w/v) SDS 50 mM TEAB (pH=7.5) and 5 mM DTT. Proteins were extracted by incubating samples at 95°C for 30 min for two cycles with 10 min sonication between cycles. Cysteine thiol side chains underwent alkylation via iodoacetamide (10 mM for 45 min at room temperature in the dark). Subsequently proteins were precipitated, purified and digested according to S-Trap procedure^14^, summarized as follows. Samples underwent acidification with 12% phosphoric acid. Proteins were precipitated by 6 times dilution with S-Trap buffer (100 mM TEAB in 90% MeOH), and then captured on porous micro S-Trap spin column beads, which were in turn washed thrice with 150 ul of S-Trap buffer. Digestion was performed in 15 ul of 50 mM ammonium bicarbonate with 400 ng trypsin (Promega) for 1 h at 47°C. Peptides were then eluted and desalted on C18 StageTips.

### LC-MS/MS

EASY-nLC 1000 (Thermo Scientific) was utilized for LC separation with precolumn Acclaim PepMap 100 (75 um x 2 cm) and analytical column PepMap RSLC C18 (2 μm, 100A x 50 cm). From the column, peptides underwent gradient elution to an Orbitrap HFX mass spectrometer via a 160 min ACN gradient from 5 to 26 % B in 118 min, then by ramp to 40% B in 20 min, and then equilibration in 100% B for 15 min (A=2% ACN 0.5% AcOH / B=80% ACN 0.5% AcOH).

Flowrate was 200 nl/min. High resolution full MS spectra were obtained with 120,000 resolution, 3e6 AGC target, 100ms maximum ion injection time, and 400-1600 m/z scan range. After every full MS scan, twenty data-dependent HCD MS/MS scans were acquired at 30,000 resolution, 5e5 AGC target, 100ms maximum ion time, one microscan, isolation window of 1.4 m/z, 27 nce, and 45 s of dynamic exclusion.

### Proteomics data analysis

Analysis of MS data was performed utilizing MaxQuant (v1.6.3.4)^15^, with the embedded Andromeda^16^ used to perform searches against the human uniprot database’s (http://www.uniprot.org/) SwissProt subset, containing 20,431 entries, augmented with 248 common laboratory contaminants and all reversed sequences. Searching parameters included enzyme specificity equal to trypsin, maximum number of missed cleavages of 2, and precursor mass tolerance of 20 ppm for the first search used for non-linear mass re-calibration^17^, and for the main search adjusted to 6 ppm. Variable modification included methionine oxidation. Fixed modification included cysteine carbamidomethylation. Minimum peptide length was 6. False discovery rate (FDR) for identifications was 1%. The option for ‘match between runs’ was utilized with a one minute retention time window. MaxQuant LFQ algorithm^18^ was used for protein quantification with minimum ratio count set to 2, “fastLFQ” option, and minimum/average number of neighbors set to 3/6. Additional analyses were done with Perseus^19^ (http://www.perseus-framework.org/) or R statistics and graphics (http://www.r-project.org/).

## Results

We first explored differences in AD pathology distribution across CA1 sPN and dPN sublayers. Using the NYU Alzheimer’s Disease Center Brain Bank, we evaluated AD pathology in a series of Braak V-VI AD brains (n = 10), with characteristics presented in Table 1, in which the Bielschowsky stain to visualize amyloid plaques and NFTs in sPN and dPN layers in both distal and proximal CA1 (Figure 1AB). Plaque density was significantly higher in the sPN layer (Figure 1C), more than 2-fold in distal CA1 near subiculum (LEC-innervated zone) and 4-fold in proximal near CA2 (MEC-innervated zone). This is consistent with qualitative observations by others^20^. We also quantified the percent of cells harboring NFTs in each layer by normalizing NFT counts by cell counts (H&E staining). The overall range of NFT propensity was similar to other work using the same approach^21^. However, analyzing the layers separately (Figure 1D) revealed that sPNs were over twice as likely to harbor NFTs in both distal and proximal CA1. Thus, sPNs and the sPN layer are more susceptible to developing NFTs and plaque, in both the LEC- and MEC-target zones of CA1.

**Table 1:** Characteristics of cases used for amyloid plaque and NFT analyses.

| Age Range | Total (N) | Female (N) | Braak Stage V (N) | Braak Stage VI (N) |
| --- | --- | --- | --- | --- |
| 61-70 | 1 | 0 | 0 | 1 |
| 71-80 | 3 | 0 | 0 | 3 |
| 81-90 | 6 | 3 | 1 | 5 |

**Figure 1.**
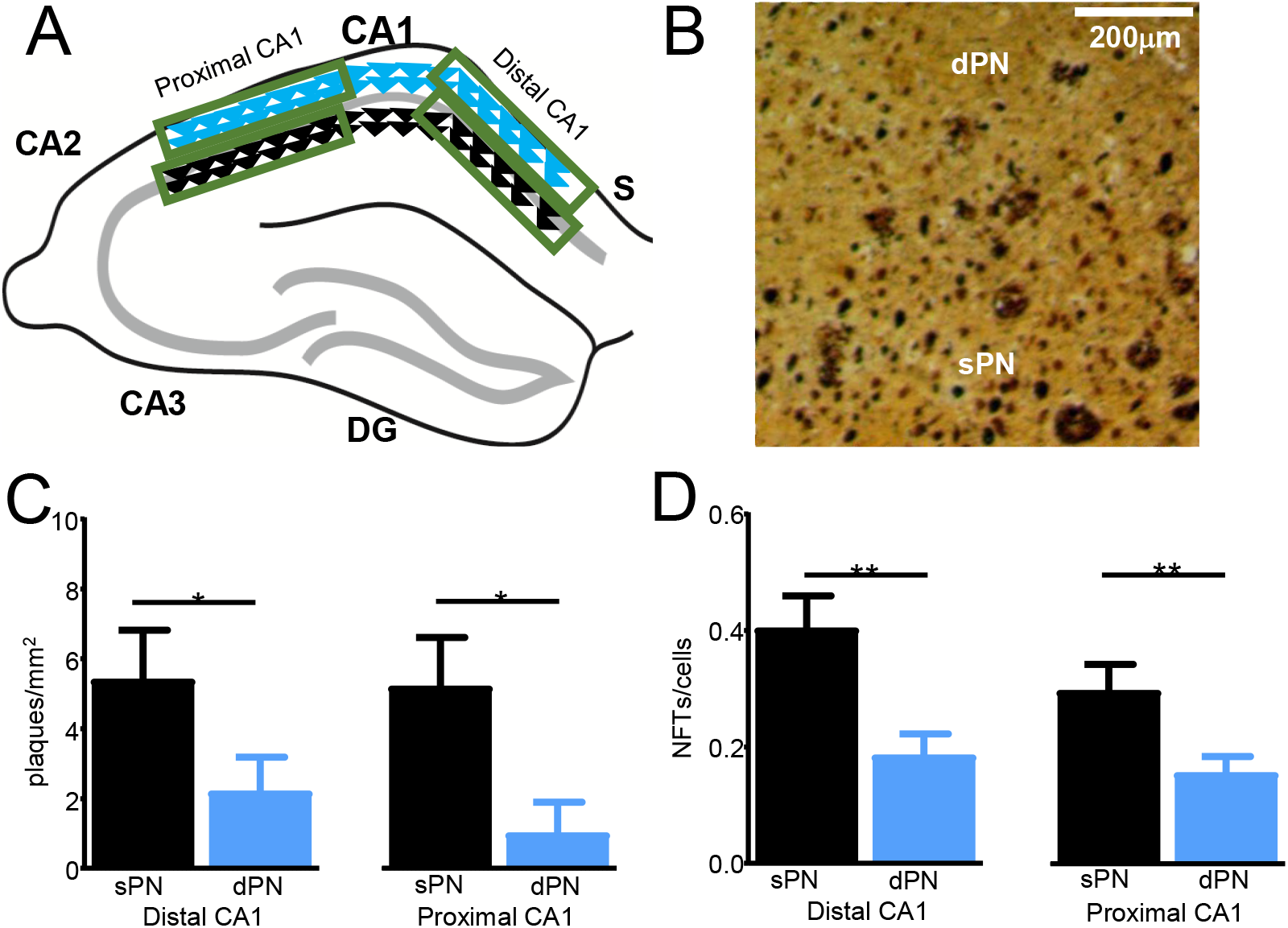
Increased AD pathology in human CA1 sPN layer. **A**. Scheme showing sPN and dPN layers in distal and proximal CA1. **B**. Bielschowsky stain of human CA1 with Braak VI AD. Small intraneuronal inclusions = NFTs, large deposits = amyloid plaques. dPN layer is above, sPN layer is below. From n = 10 Braak VI AD brains, quantification of **B**. Plaque density and **C**. Frequency of NFTs per cell (=NFT density/cell density via H&E stain). *p<0.002; **p<0.0001.

We next analyzed the proteome of human CA1 sPN/dPN layers in a separate set of advanced AD brains (n = 8). Characteristics of these case are presented in Table 2. Our analyses focused on PNs in distal CA1, given that the burden of AD pathology is higher there (Figure 2AB). Overall in our analyses we were able to detect and quantify 3159 proteins from LCM-dissected neurons using FFPE samples, spanning ∼6 orders of magnitude in abundance, and with robust capture (∼17% from all identifications) of 537 synapse and ion channel-related proteins (Figure 2CD). Despite this small group, we identified several differences (Figure 3, labeled) that included peptides to cell health, physiologic function, or disease. These findings are summarized in Table 3. Most hits showed relative elevations in dPNs. Significant hits related to AMPAR regulation (*SIRT2*) ^22^, synapse function (*ARHGAP23, SEPTIN4*) ^23,24^, markers of neurodegeneration (*NEFH*) ^25^, and markers of late versus early AD vulnerability (*INA*) ^26^. Nearly significant hits included potassium channel-associated *CNTN2* ^27,28^, postsynaptic density proteins *DLG4* and variants, which are dysregulated in AD ^29^, and GABAB modulator *KCTD16* ^30^. These results suggested that even in the advanced AD brain, human sPN and dPN layers show molecular differences that relate to physiological function and neurodegeneration.

**Table 2:** Characteristics of cases used for proteomic analyses.

| Age Range | Total (N) | Female (N) | Braak Stage VI (N) |
| --- | --- | --- | --- |
| 70-79 | 3 | 0 | 3 |
| 80-89 | 2 | 2 | 2 |
| 90-99 | 3 | 2 | 3 |

**Table 3:**
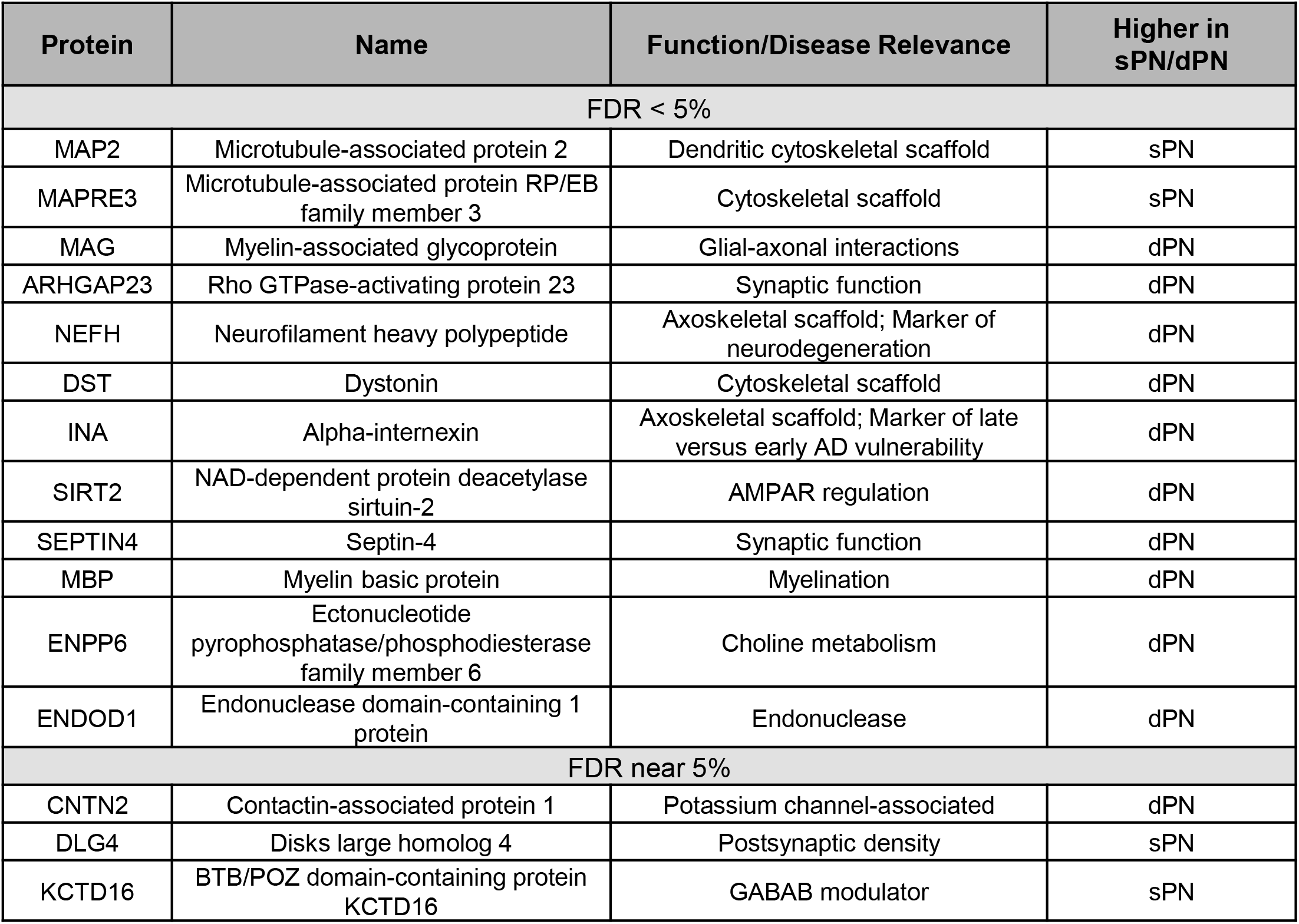
Significant and near-significant hits from proteomic analysis.

**Figure 2.**
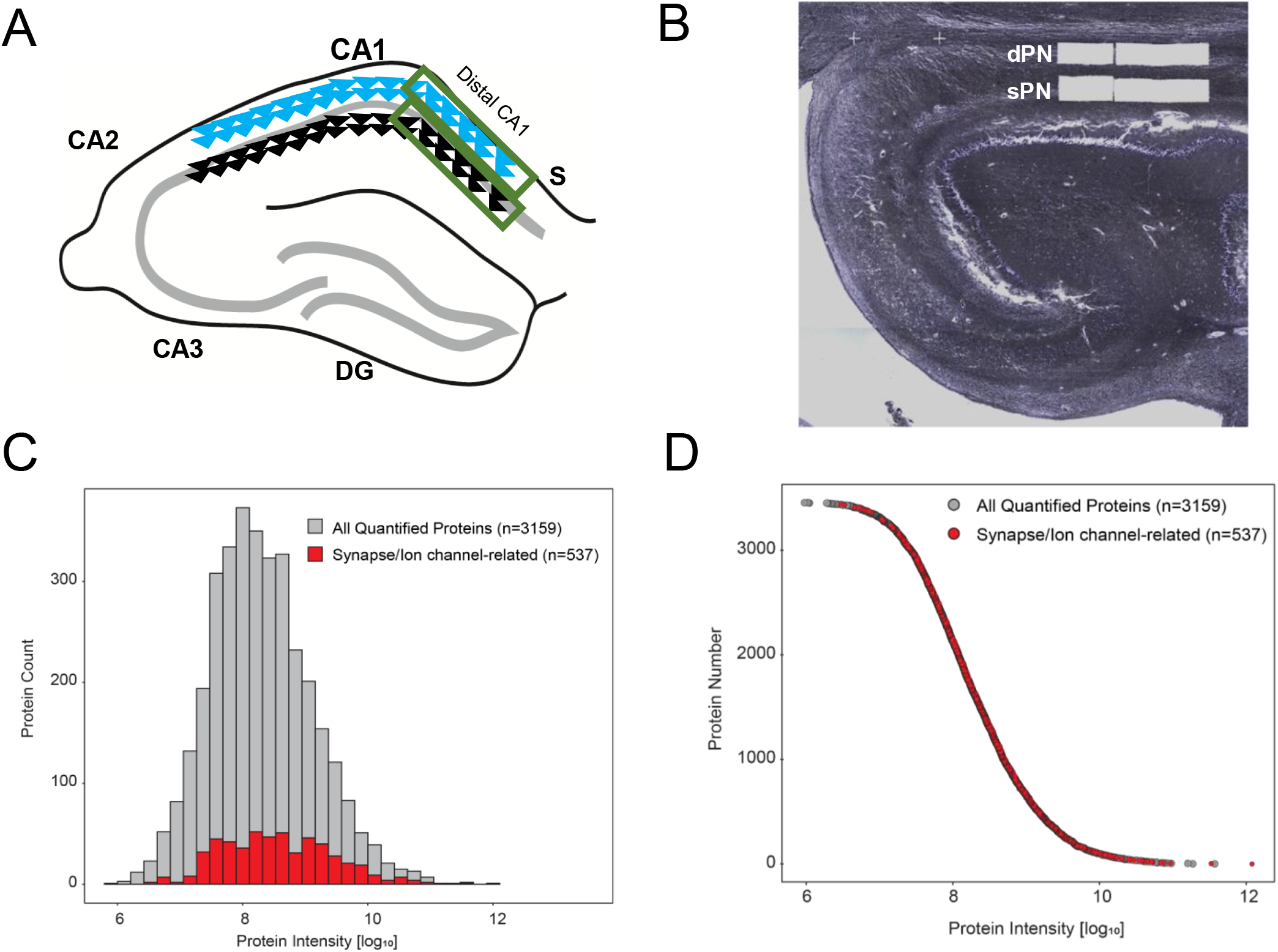
Quantitative, localized proteomics of human CA1 sPN and dPN layers in AD. **A**. Scheme showing focus of analysis on sPNs and dPNs of distal CA1. **B**. Light microscopic image of human CA1 section and sPN and dPN layers extracted via laser capture microdissection. Yield of proteomic analysis of sPNs and dPNs of n = 8 AD brains demonstrated as follow. **C**. Proteins related to synapses and ion channels are in red. Over 3000 unique proteins were identified, spanning 6 orders of magnitude. There was excellent coverage of synapse- and ion-channel related protein (∼15% of identifications). **D**. Alternative view of data in C, showing individual synapse- and ion-channel related proteins (red) are distributed in a full range of intensities

**Figure 3.**
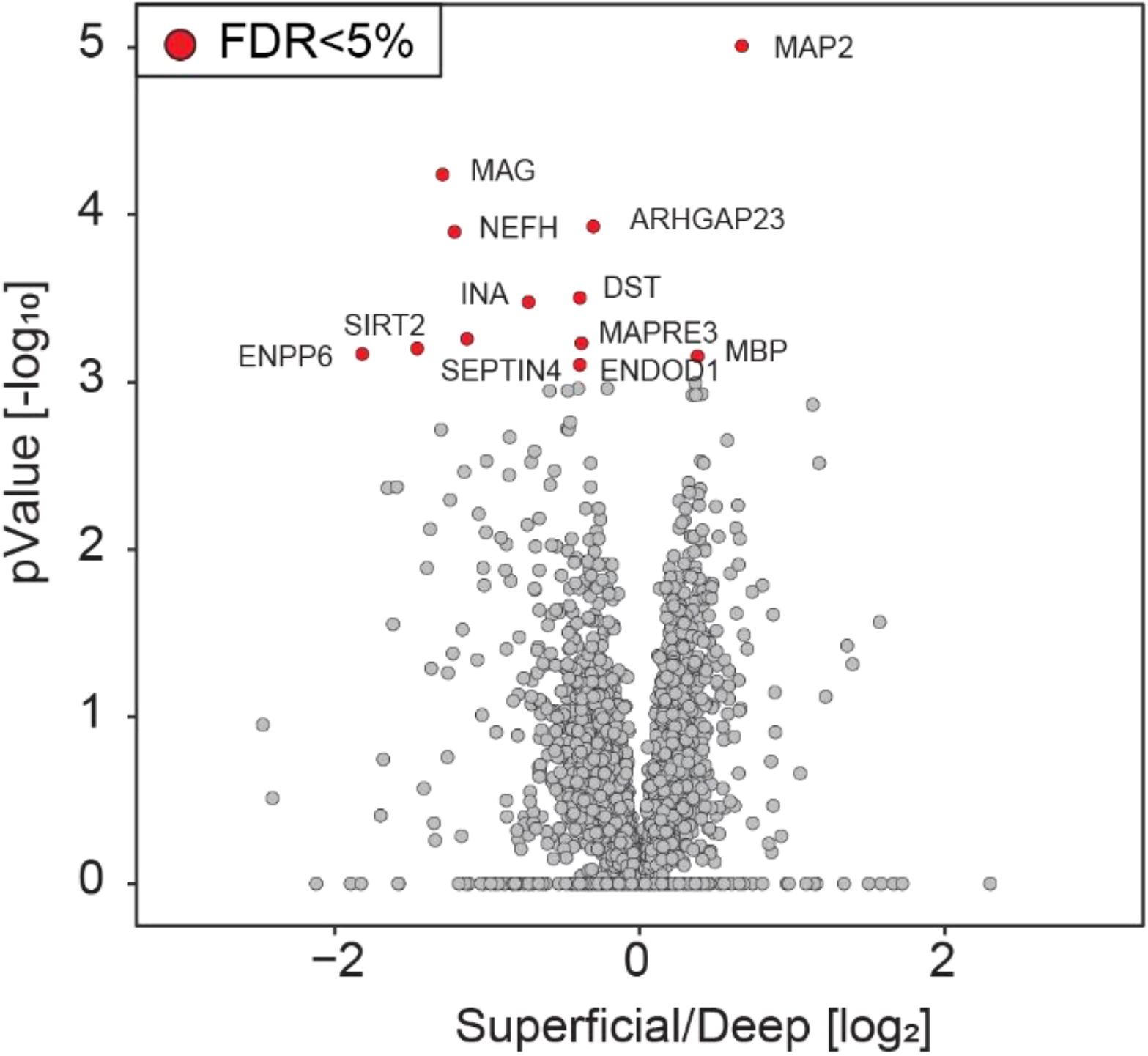
Volcano plot of CA1 sPN, dPN proteomes in human AD. Two sample paired t-test was used to identify protein hits which abundance is significantly different (permutation-based FDR correction, FDR cutoff at 5%) between CA1 sPNs and dPNs (n = 8/group). Statistically significant hits are identified in red with gene name indicated.

To probe deeper into potential mechanisms of the sPN layer’s differential vulnerability to develop AD pathology, we examined a marker of excitatory-inhibitory (E/I) balance, the DLG4/GPHN ratio, which relates to postsynaptic markers of glutamatergic and GABAergic synapses^31,32^. In 7 of the 8 cases, sPNs displayed a higher DLG4/GPHN ratio (Figure 4A). To examine if this was driven by DLG4 or GPHN, we performed a linear correlation analysis. This demonstrated that the elevated DLG4/GPHN ratio was due to DLG4, as it showed a positive linear correlation with this ratio, whereas GPHN did not show a negative linear correlation (Figure 4BC). In sum, these results suggested that sPNs show relative hyperexcitability compared to dPNs in AD, possibly driven by postsynaptic glutamatergic differences.

**Figure 4.**
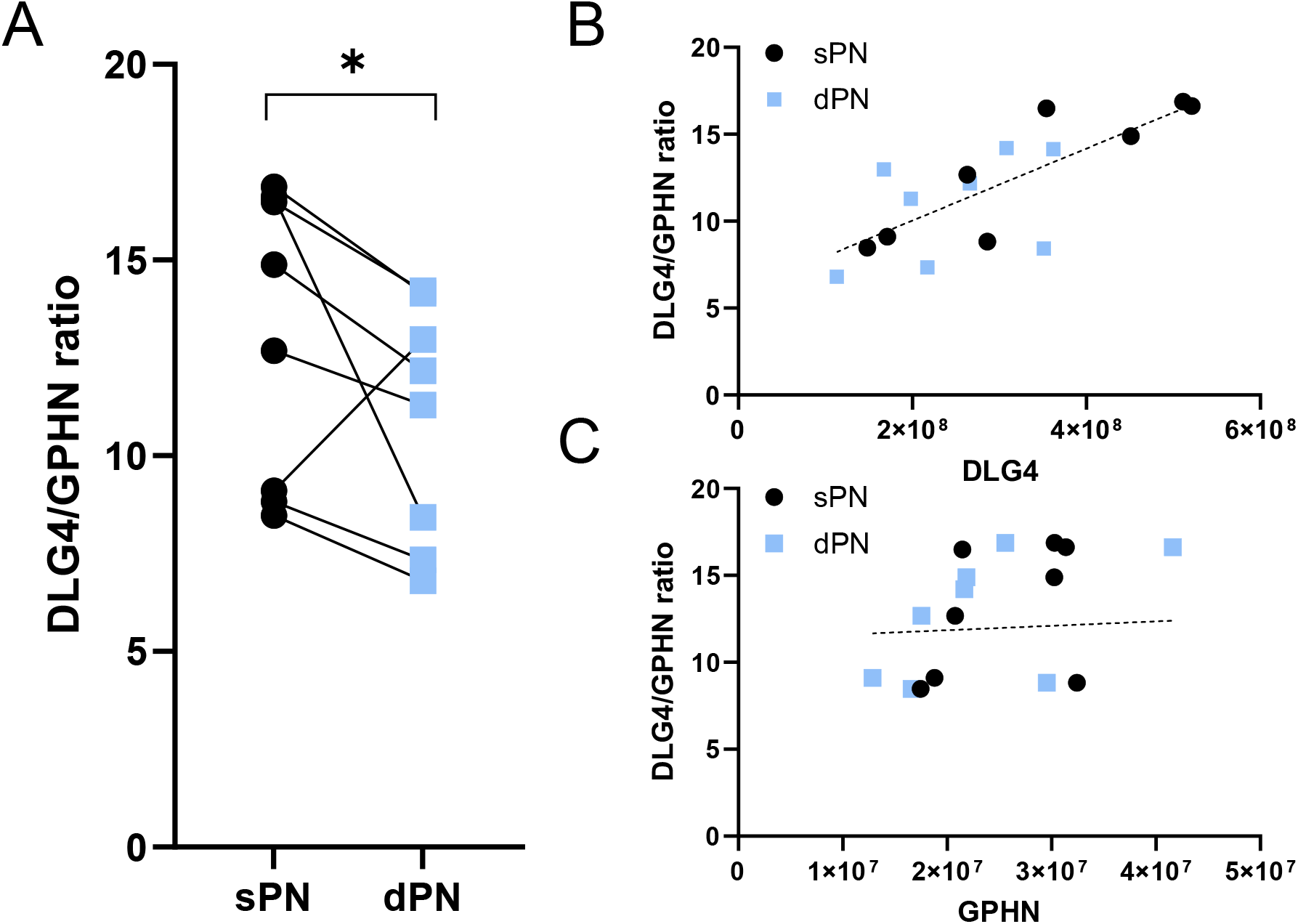
Increased hyperexcitability marker DLG4/GPHN ratio in sPNs in human AD. **A**. In 7/8 cases, CA1 sPNs featured a higher DLG4/GPHN ratio than CA1 dPNs (p = 0.01 by Fisher’s exact test). Linear correlation analysis revealed that this was driven by **B**. positive correlation with DLG4 (p = 0.0007, R^2^ = 0.5744) and not by **C**. negative correlation with GPHN (p = 0.84, R^2^ = 0.003).

## Discussion

In this study we found that in advanced human AD brains, sPN and dPN layers show differential patterns of AD pathology severity, functional- and neurodegeneration-related molecular profiles, and E/I balance. Of note, the sPN layer shows a higher propensity of amyloid and tau pathology, regardless of cortical innervation zone. This suggests that intrinsic vulnerability may play a role, since cortical drive shifts from sPNs to dPNs across the distal-proximal axis ^4^.

Consistent with this, sPNs show higher postsynaptic markers of E/I balance that would implicate relative hyperexcitability as a contributing mechanism, as it has been implicated in spread of both amyloid and tau pathology ^33^. Of note, analysis of rodent circuity has demonstrated that sPNs are indeed prone to hyperexcitability relative to dPNs related to differences in excitatory response integration as well as inhibition ^4,6,34^. While our human proteomic analyses did not reveal these differences, molecular analysis of sPNs in rodents also show that sPNs show higher levels of c-jun N-terminal kinase ^35^ and the 5-HT1A receptor ^36^, both of which have been shown to enhance AD pathogenesis ^37,38^. Mirroring this, our proteomic analysis also revealed that dPNs showed relatively higher levels of neurofilament heavy polypeptide (*NEFH*) a marker of active neurodegeneration ^25^, and alpha-internexin (*INA*), a proposed marker of hierarchical vulnerability to AD and specifically of later, as opposed to earlier, degeneration in AD ^26^. In sum, this may suggest that sPNs are vulnerable in earlier stages, with increased development of AD pathology in that layer, whereas dPNs are somewhat protected early on and become involved in a delayed fashion in later stage AD.

We acknowledge that our study has some limitations that would instruct future work. First, our sample size was small and analysis of a larger cohort may result in more proteomic hits that would enhance pathway analysis. Second, we were not able to compare to control cases to determine if sPNs and dPNs were changing differentially with respect to those populations in brain without AD pathology. Lastly, it will be important to analyze pathology development and proteomic changes in early stage AD to verify the working hypothesis established by these findings that sPNs are vulnerable early, before dPNs.

## Data Availability

All data produced in the present study are available upon reasonable request to the authors

## Acknowledgements

This work was funded by NIH P30AG008051 and P30AG066512.

## References

1 Braak, H. & Braak, E. Neuropathological stageing of Alzheimer-related changes. Acta Neuropathol 82, 239–259 (1991).

2 Lace, G. et al. Hippocampal tau pathology is related to neuroanatomical connections: an ageing population-based study. Brain 132, 1324–1334, doi:10.1093/brain/awp059 (2009).

3 Masurkar, A. V. Towards a circuit-level understanding of hippocampal CA1 dysfunction in Alzheimer’s disease across anatomical axes. J Alzheimers Dis Parkinsonism 8 (2018).

4 Masurkar, A. V. et al. Medial and Lateral Entorhinal Cortex Differentially Excite Deep versus Superficial CA1 Pyramidal Neurons. Cell Rep 18, 148–160, doi:10.1016/j.celrep.2016.12.012 (2017).

5 Khan, U. A. et al. Molecular drivers and cortical spread of lateral entorhinal cortex dysfunction in preclinical Alzheimer’s disease. Nat Neurosci 17, 304–311, doi:10.1038/nn.3606 (2014).

6 Masurkar, A. V. et al. Postsynaptic integrative properties of dorsal CA1 pyramidal neuron subpopulations. J Neurophysiol 123, 980–992, doi:10.1152/jn.00397.2019 (2020).

7 Hondius, D. C. et al. Profiling the human hippocampal proteome at all pathologic stages of Alzheimer’s disease. Alzheimers Dement 12, 654–668, doi:10.1016/j.jalz.2015.11.002 (2016).

8 Schrotter, A. et al. LMD proteomics provides evidence for hippocampus field-specific motor protein abundance changes with relevance to Alzheimer’s disease. Biochim Biophys Acta Proteins Proteom 1865, 703–714, doi:10.1016/j.bbapap.2017.03.013 (2017).

9 Wang, Q. et al. Proteomic analysis of neurofibrillary tangles in Alzheimer disease identifies GAPDH as a detergent-insoluble paired helical filament tau binding protein. FASEB J 19, 869–871, doi:10.1096/fj.04-3210fje (2005).

10 Drummond, E. S., Nayak, S., Ueberheide, B. & Wisniewski, T. Proteomic analysis of neurons microdissected from formalin-fixed, paraffin-embedded Alzheimer’s disease brain tissue. Sci Rep 5, 15456, doi:10.1038/srep15456 (2015).

11 Drummond, E. et al. Proteomic differences in amyloid plaques in rapidly progressive and sporadic Alzheimer’s disease. Acta Neuropathol 133, 933–954, doi:10.1007/s00401-017-1691-0 (2017).

12 Drummond, E. & Wisniewski, T. The use of localized proteomics to identify the drivers of Alzheimer’s disease pathogenesis. Neural Regen Res 12, 912–913, doi:10.4103/1673-5374.208570 (2017).

13 Drummond, E., Nayak, S., Pires, G., Ueberheide, B. & Wisniewski, T. Isolation of Amyloid Plaques and Neurofibrillary Tangles from Archived Alzheimer’s Disease Tissue Using Laser-Capture Microdissection for Downstream Proteomics. Methods Mol Biol 1723, 319–334, doi:10.1007/978-1-4939-7558-7_18 (2018).

14 HaileMariam, M. et al. S-Trap, an Ultrafast Sample-Preparation Approach for Shotgun Proteomics. J Proteome Res 17, 2917–2924, doi:10.1021/acs.jproteome.8b00505 (2018).

15 Cox, J. & Mann, M. MaxQuant enables high peptide identification rates, individualized p.p.b.-range mass accuracies and proteome-wide protein quantification. Nat Biotechnol 26, 1367–1372, doi:10.1038/nbt.1511 (2008).

16 Cox, J. et al. Andromeda: a peptide search engine integrated into the MaxQuant environment. J Proteome Res 10, 1794–1805, doi:10.1021/pr101065j (2011).

17 Cox, J., Michalski, A. & Mann, M. Software lock mass by two-dimensional minimization of peptide mass errors. J Am Soc Mass Spectrom 22, 1373–1380, doi:10.1007/s13361-011-0142-8 (2011).

18 Cox, J. et al. Accurate proteome-wide label-free quantification by delayed normalization and maximal peptide ratio extraction, termed MaxLFQ. Mol Cell Proteomics 13, 2513–2526, doi:10.1074/mcp.M113.031591 (2014).

19 Tyanova, S. et al. The Perseus computational platform for comprehensive analysis of (prote)omics data. Nat Methods 13, 731–740, doi:10.1038/nmeth.3901 (2016).

20 Llorens-Martin, M. et al. Selective alterations of neurons and circuits related to early memory loss in Alzheimer’s disease. Front Neuroanat 8, 38, doi:10.3389/fnana.2014.00038 (2014).

21 Hof, P. R. et al. Stereologic evidence for persistence of viable neurons in layer II of the entorhinal cortex and the CA1 field in Alzheimer disease. J Neuropathol Exp Neurol 62, 55–67, doi:10.1093/jnen/62.1.55 (2003).

22 Wang, G. et al. Crucial Roles for SIRT2 and AMPA Receptor Acetylation in Synaptic Plasticity and Memory. Cell Rep 20, 1335–1347, doi:10.1016/j.celrep.2017.07.030 (2017).

23 Martin-Vilchez, S. et al. RhoGTPase Regulators Orchestrate Distinct Stages of Synaptic Development. PLoS One 12, e0170464, doi:10.1371/journal.pone.0170464 (2017).

24 Marttinen, M., Kurkinen, K. M., Soininen, H., Haapasalo, A. & Hiltunen, M. Synaptic dysfunction and septin protein family members in neurodegenerative diseases. Mol Neurodegener 10, 16, doi:10.1186/s13024-015-0013-z (2015).

25 Noh, K. M. et al. Repressor element-1 silencing transcription factor (REST)-dependent epigenetic remodeling is critical to ischemia-induced neuronal death. Proc Natl Acad Sci U S A 109, E962–971, doi:10.1073/pnas.1121568109 (2012).

26 Dickson, T. C., Chuckowree, J. A., Chuah, M. I., West, A. K. & Vickers, J. C. alpha-Internexin immunoreactivity reflects variable neuronal vulnerability in Alzheimer’s disease and supports the role of the beta-amyloid plaques in inducing neuronal injury. Neurobiol Dis 18, 286–295, doi:10.1016/j.nbd.2004.10.001 (2005).

27 Gu, C. & Gu, Y. Clustering and activity tuning of Kv1 channels in myelinated hippocampal axons. J Biol Chem 286, 25835–25847, doi:10.1074/jbc.M111.219113 (2011).

28 Pinatel, D. et al. The Kv1-associated molecules TAG-1 and Caspr2 are selectively targeted to the axon initial segment in hippocampal neurons. J Cell Sci 130, 2209–2220, doi:10.1242/jcs.202267 (2017).

29 Savioz, A., Leuba, G. & Vallet, P. G. A framework to understand the variations of PSD-95 expression in brain aging and in Alzheimer’s disease. Ageing Res Rev 18, 86–94, doi:10.1016/j.arr.2014.09.004 (2014).

30 Fritzius, T. et al. KCTD Hetero-oligomers Confer Unique Kinetic Properties on Hippocampal GABAB Receptor-Induced K+ Currents. J Neurosci 37, 1162–1175, doi:10.1523/JNEUROSCI.2181-16.2016 (2017).

31 Lauterborn, J. C. et al. Increased excitatory to inhibitory synaptic ratio in parietal cortex samples from individuals with Alzheimer’s disease. Nat Commun 12, 2603, doi:10.1038/s41467-021-22742-8 (2021).

32 Scaduto, P. et al. Functional excitatory to inhibitory synaptic imbalance and loss of cognitive performance in people with Alzheimer’s disease neuropathologic change. Acta Neuropathol 145, 303–324, doi:10.1007/s00401-022-02526-0 (2023).

33 Kamondi, A., Grigg-Damberger, M., Loscher, W., Tanila, H. & Horvath, A. A. Epilepsy and epileptiform activity in late-onset Alzheimer disease: clinical and pathophysiological advances, gaps and conundrums. Nat Rev Neurol 20, 162–182, doi:10.1038/s41582-024-00932-4 (2024).

34 Lee, S. H. et al. Parvalbumin-positive basket cells differentiate among hippocampal pyramidal cells. Neuron 82, 1129–1144, doi:10.1016/j.neuron.2014.03.034 (2014).

35 Maroso, M. et al. Cannabinoid Control of Learning and Memory through HCN Channels. Neuron 89, 1059–1073, doi:10.1016/j.neuron.2016.01.023 (2016).

36 Cembrowski, M. S. et al. Spatial Gene-Expression Gradients Underlie Prominent Heterogeneity of CA1 Pyramidal Neurons. Neuron 89, 351–368, doi:10.1016/j.neuron.2015.12.013 (2016).

37 Wang, Y. J. et al. Escitalopram attenuates beta-amyloid-induced tau hyperphosphorylation in primary hippocampal neurons through the 5-HT1A receptor mediated Akt/GSK-3beta pathway. Oncotarget 7, 13328–13339, doi:10.18632/oncotarget.7798 (2016).

38 Yarza, R., Vela, S., Solas, M. & Ramirez, M. J. c-Jun N-terminal Kinase (JNK) Signaling as a Therapeutic Target for Alzheimer’s Disease. Front Pharmacol 6, 321, doi:10.3389/fphar.2015.00321 (2015).

